# The time-varying association between previous antibiotic use and antibiotic resistance

**DOI:** 10.1101/2022.08.30.22279375

**Authors:** Avi Baraz, Michal Chowers, Daniel Nevo, Uri Obolski

## Abstract

**Objectives:** The objective of the study was to estimate how the time elapsed from previous antibiotic use is associated with antibiotic resistance.

**Methods:** Data comprised electronic medical records of all patients in an Israeli hospital who had a positive bacterial culture between 2016-19. These included susceptibility testing results, and clinical and demographic data. Mixed-effects time-varying logistic models were fitted to estimate the association between time elapsed since last use of aminoglycosides and gentamicin resistance (*n* = 13,095), cephalosporins and ceftazidime resistance (*n* = 13,051), and fluoroquinolones and ciprofloxacin resistance (*n* = 15,364), while adjusting for multiple covariates.

**Results:** For all examined antibiotics, previous antibiotic use had a statistically significant association with resistance (*p* < 0.001). These associations exhibited a clear decreasing pattern over time, which we present as a flexible function of time. Nonetheless, previous antibiotic use remained a significant risk factor for resistance for at least 180 days, with adjusted odds ratios of 1.94 (95% CI 1.40-2.69), 1.33 (95% CI 1.10-1.61), and 2.25 (95% CI 1.49-3.41), for gentamicin, ceftazidime and ciprofloxacin, respectively.

**Conclusions:** The association between prior antibiotic use and resistance decreases over time. Commonly used cut-offs for prior antibiotic use can either misclassify patients still at higher risk, when too recent; or provide a diluted estimate of the effects of antibiotic use on future resistance, when too distant. Hence, prior antibiotic use should be considered as a time-dependent risk factor for resistance, both in epidemiological research and clinical practice.

## Introduction

Empiric antibiotic therapy is a common and crucial component of treating hospitalized patients. However, increased antibiotic resistance frequencies often lead to inappropriate empiric therapy [1]. Risk factors for antibiotic resistant infections are hence constantly investigated, for both research and practical, clinical purposes [2]. A biologically plausible and empirically evident cause for resistance is previous use of antibiotics [3]. Despite its importance, little is known about the association between the time elapsed since antibiotic use on the risk for resistance. Both prospective and retrospective studies ubiquitously report previous antibiotic use as a binary variable, indicating prior antibiotic use in certain, often arbitrary time-periods, with common cut-offs of 30, 90, or 365 days [2,4-11]. Such discretization of the data can disregard patients with existing risk if the cut-off is too short, or dilute the effects of the risk if the cut-off is too long. Here, we employed patients’ electronic medical records (EMRs) to estimate the risk of antibiotic resistance as a function of the time elapsed since their last antibiotic use. We applied mixed-effects time-varying logistic regression and obtained the adjusted risk estimates as flexible functions of time elapsed since last use of antibiotics for gentamicin, ceftazidime and ciprofloxacin.

## Materials and methods

### Data

The dataset included the EMRs of all patients who had a positive bacterial culture between 2016-19, in Meir medical center, Israel. The data contained, patients’ antibiotic susceptibility test results from all positive cultures, as well as demographic and clinical data, including antibiotic use within the hospital in the year preceding the susceptibility tests (see Supplementary information, Tables S1-S3). Cultures were taken upon suspected infection, but were not verified against clinical symptoms.

The main variable of interest was the time elapsed (in days) since a patients’ last antibiotic use in the hospital until the drawing of their bacterial cultures. Antibiotic use was considered only when it commenced ≥ 5 days and ≤ 365 days before a culture was drawn. The first constraint was set as to avoid date misclassifications that might lead to antibiotics prescribed after culture results were obtained, and since it is a short period for resistance acquisition. The second constraint was set due to data availability. To attain a sufficient sample size, antibiotic use was grouped into cephalosporins (of all generations), fluoroquinolones and aminoglycosides, administered either orally or intravenously. Resistance to three antibiotics, selected as markers for resistance to the classes of antibiotics considered above, was modeled: gentamicin, ceftazidime, and ciprofloxacin.

### Statistical analysis

Relationships between patients’ clinical and demographic covariates, and antibiotic resistance were modeled by mixed-effects time-varying logistic models [12]. These models extend standard logistic regression by allowing the effect of previous antibiotic use to vary as flexible functions of time, using splines (see Supplementary information for additional information). The models addressed multiple cultures from the same patients by assigning a random intercept per patient, although this addition did not substantially change the results. The models were adjusted for multiple prominent clinical and demographic covariates, chosen based on prior knowledge [2], and underwent sensitivity and subgroup analyses (see Supplementary information, Tables S1-S3).

Additionally, we examined adjustments for prior use of the other classes of antibiotics modeled, as well as to non-cephalosporin beta-lactam antibiotics, to account for potential cross-resistance [13]. Each of these covariates was modeled as a dummy variable (used/not), with a linear time trend (in the log-odds scale). These variables had little influence on the results, and hence only prior use of non-cephalosporin beta-lactams was incorporated into the ceftazidime resistance model (see Supplementary information, Figures S1-S3).

## Ethics declaration

The study was approved by the Institutional Review Board Committee of the Meir Medical Center (MMC-0143-19).

## Results

We examined the association between previous aminoglycoside use and resistance to gentamicin (*n* = 13,095) cultures, resistance frequency 15.58%); previous cephalosporin use and resistance to ceftazidime (*n* = 13,051 cultures, resistance frequency 24.27%); and previous fluoroquinolone use and resistance to ciprofloxacin (*n* = 15,364) cultures, resistance frequency 26.66%). For all examined antibiotics, the time-varying coefficient of previous antibiotic use in the hospital had a statistically significant association with resistance (*p* < 0.001) (see Supplementary information, Additional Information, Tables S7-S9). As expected, the spline-based adjusted odds ratios (aORs) decayed as time passed from last antibiotic use (Figure 1).

**Fig. 1.**
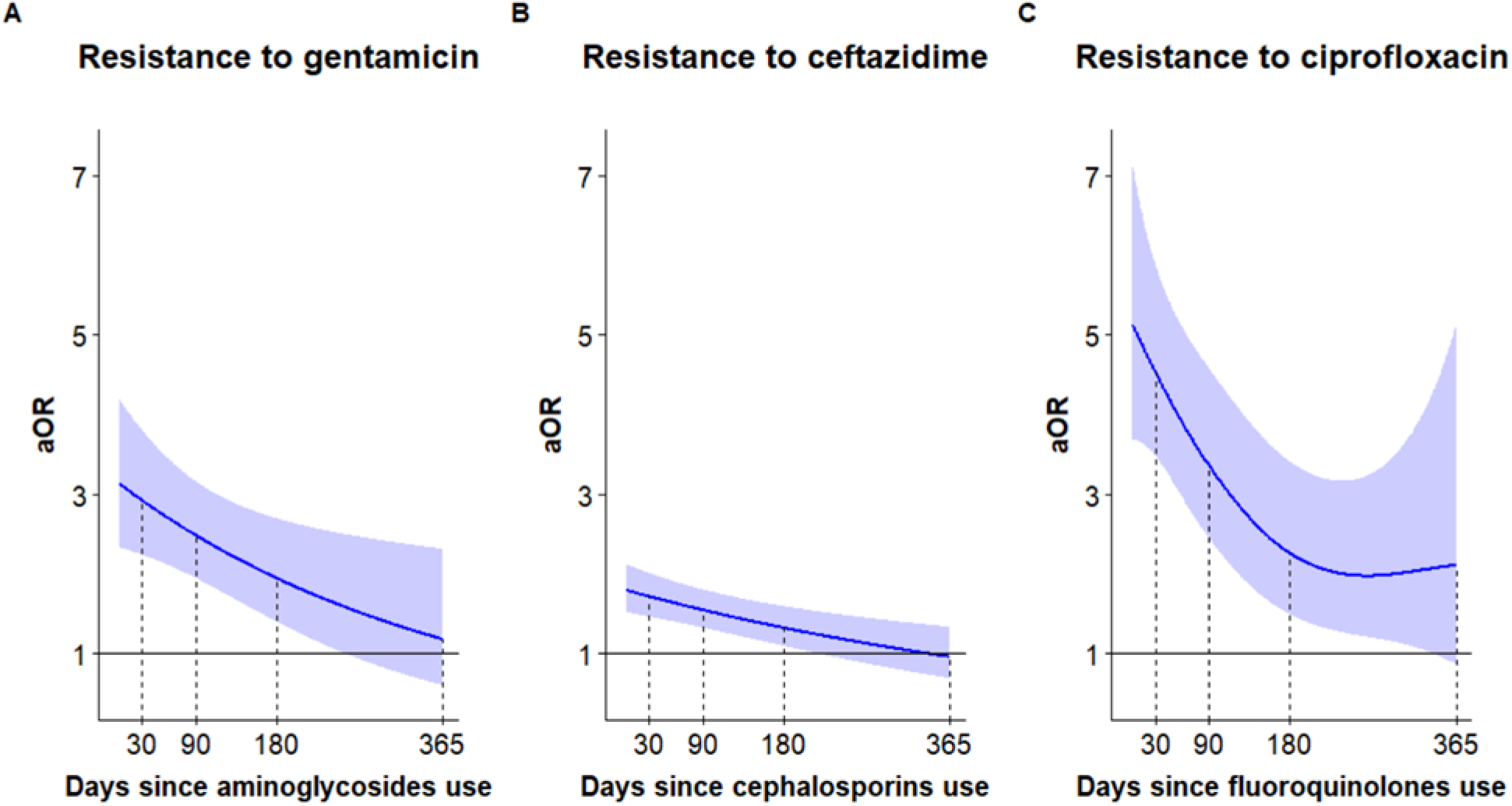
Estimates of risk for antibiotic resistance from generalized additive mixed-effects models: Spline-based estimates (curves) and their 95% CI (shaded) of the aORs for resistance, against days elapsed since last use of relevant antibiotics. (a) gentamicin resistance against last use of aminoglycosides; (b) ceftazidime resistance against last use of cephalosporins; and (c) ciprofloxacin resistance against last use of fluoroquinolones.

Importantly, the estimates across the three antibiotics examined maintained significant and positive associations between previous use and resistance for over 180 days. Summary of the aORs and their CIs at 30, 90, 180 and 365 days are provided in Table 1. For gentamicin and ceftazidime, the decrease of the aOR can be very well approximated by exponential decay of the forms and, respectively, where is months since last antibiotic use (see Supplementary material for details). The decrease in the aOR of ciprofloxacin resistance did not follow such a form, and the estimate varied substantially as time elapsed from the last use increased, possibly due to limited sample size for distant periods (Figure 1C, shaded region).

**Table 1.**
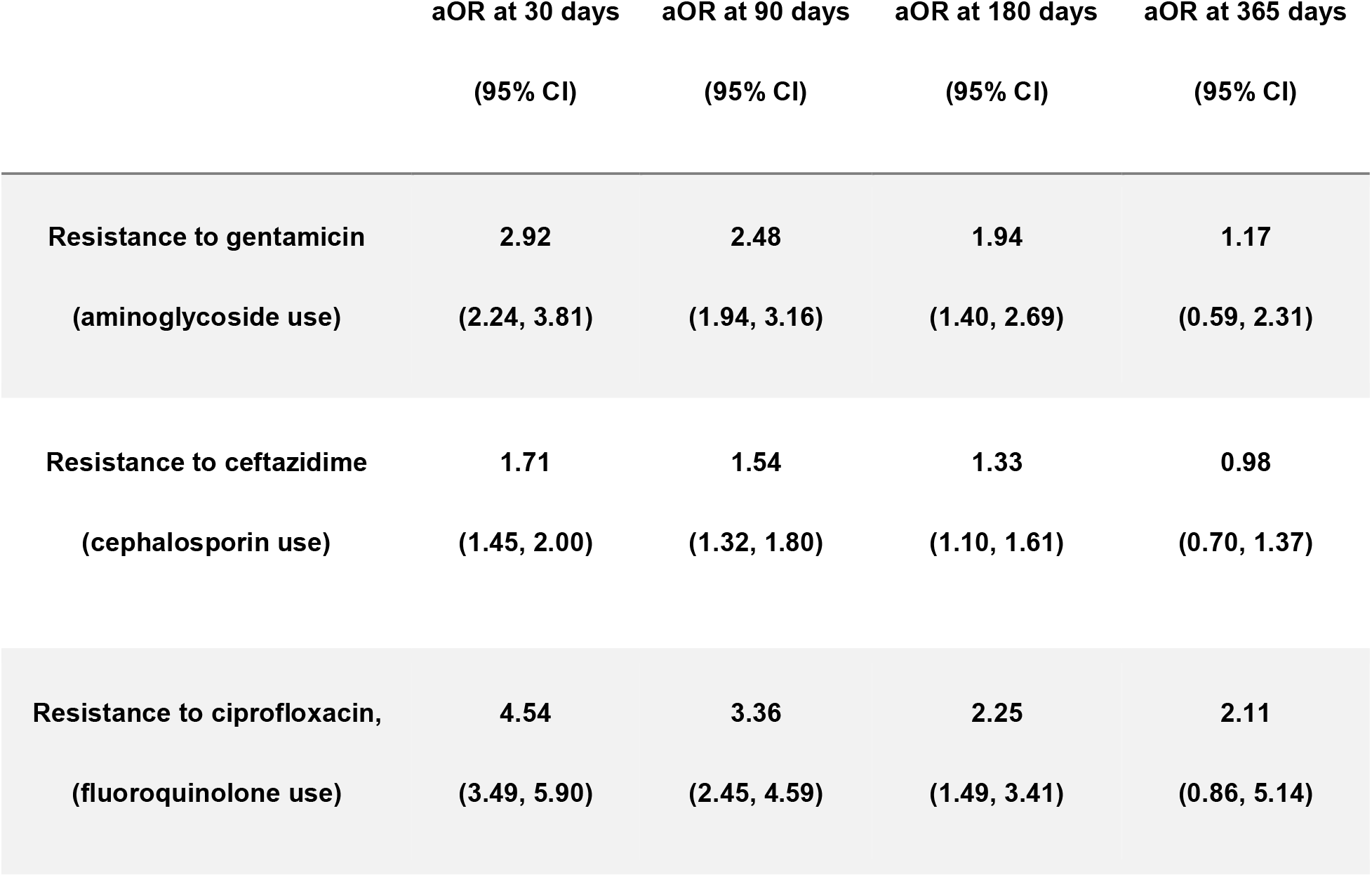
Adjusted odds-ratios (aOR) for resistance to different antibiotics by time since last antibiotic use. The aOR were adjusted for patients’ clinical and demographic characteristics, as well as to bacterial species and sample source (see Supplementary information, Tables S7-S9).

|  | aOR at 30 days<br>(95% CI) | aOR at 90 days<br>(95% CI) | aOR at 180 days<br>(95% CI) | aOR at 365 days<br>(95% CI) |
| --- | --- | --- | --- | --- |
| **Resistance to gentamicin**<br>**(aminoglycoside use)** | **2.92**<br>**(2.24, 3.81)** | **2.48**<br>**(1.94, 3.16)** | **1.94**<br>**(1.40, 2.69)** | **1.17**<br>**(0.59, 2.31)** |
| **Resistance to ceftazidime**<br>**(cephalosporin use)** | **1.71**<br>**(1.45, 2.00)** | **1.54**<br>**(1.32, 1.80)** | **1.33**<br>**(1.10, 1.61)** | **0.98**<br>**(0.70, 1.37)** |
| **Resistance to ciprofloxacin,**<br>**(fluoroquinolone use)** | **4.54**<br>**(3.49, 5.90)** | **3.36**<br>**(2.45, 4.59)** | **2.25**<br>**(1.49, 3.41)** | **2.11**<br>**(0.86, 5.14)** |

## Discussion

In this study, we provided time-varying estimates of the association of previous antibiotic use with antibiotic resistance. We have characterized these estimates for gentamicin, ceftazidime and ciprofloxacin, serving as proxies for resistance to aminoglycosides, cephalosporins and fluoroquinolones, respectively. Whereas the estimates differed between the antibiotics examined, they all exhibited a marked decrease over time. These differences might be explained by the variation of the link between exposure and resistance acquisition between antibiotic classes [8]. Higher aORs are expected for fluoroquinolones relative to aminoglycosides [8]. However, the relatively low aORs obtained for cephalosporins might be due to the aggregation of different cephalosporins administered to patients. 1st and 2nd generation cephalosporins are more widely used in the studied hospital, and are expected to confer lower resistance to ceftazidime, compared to 3rd generation cephalosporins [8,14].

Nonetheless, for all examined antibiotics, previous antibiotic use remained a statistically significant risk factor for resistance for at least 180 days.

Our results have important implications for both research and clinical practice. Studies analyzing risk factors for antibiotic resistance model previous antibiotic use as binary indicators, choosing cut-offs ranging from several days to a year prior to the examined bacterial sample. Our results demonstrated that this might lead to inaccuracies. Applying antibiotic use cut-offs close to the outcome (e.g. 30 or 90 days), might lead to substantial underestimation of the risk of patients using antibiotics before the cut-offs. Conversely, applying distant cut-offs (e.g. one year) might lead to underestimation of the association between previous use and resistance for patients who used antibiotics recently. One solution considers the time elapsed since antibiotic use, either with splines as demonstrated here, or approximated by linear interactions with time in a standard logistic regression model. Furthermore, clinicians should bear in mind that patients who previously used antibiotics are more prone to have resistant infections, and that this risk decays over time but persists for at least 180 days.

Our study has several limitations. Despite known biological mechanisms linking antibiotic use to resistance, the results presented here do not claim to be causal estimates. Residual confounding could still affect our estimates. For example, we did not possess patient data of antibiotic use outside the hospital. This could have contributed to the high variance in the fluoroquinolone estimates or affected the cephalosporin estimates, as fluoroquinolones and older generation cephalosporins may be prescribed to outpatients. However, since our models adjust for many other variables that could account for these unobserved differences between patients, we hope that these effects are reasonably mitigated. Furthermore, the sample collection dates in our data reflect clinical infection onset, but not necessarily the patient’s microbiota. Ideally, a study prospectively sampling patients could more accurately inform on the formation of antibiotic resistant bacterial populations. Finally, this is a single center study, and the estimates presented here could vary in settings with different antibiotic resistance frequencies, patient populations, or other conditions affecting bacterial infections. Hence, although the general time-varying patterns observed here may persist, they are best estimated locally, for more accurate risk stratification of patients. Nonetheless, the implications of our estimates are relevant for studies attempting to estimate risk factors, or predictors, for resistance, regardless of causal interpretation. This is true both for epidemiological studies using these variables, or when clinicians utilize previous antibiotic use to estimate the risk of antibiotic resistant infections for hospitalized patients.

In conclusion, this study provides time-varying estimates of the association of antibiotic use with future antibiotic resistance. These estimates are relevant both for epidemiological literature and clinical practice. Commonly used cut-offs for previous use can either misclassify patients still at higher risk, or provide diluted estimates of the association of antibiotic use with future resistance. We therefore implore to consider prior antibiotic use as a time-dependent risk factor for resistance, and to continue researching this phenomenon for various antibiotics and settings.

## Data Availability

The data and computing code are not available for replication because they are protected under medical confidentiality.

## Transparency declaration

There are no potential conflicts of interest for any authors.

## Author contributions

UO and DN conceived the study and supervised all analysis; AB performed the analyses; MC provided clinical insights; All authors interpreted the results; AB, DN, and UO wrote the initial draft; all authors critically revised and approved the paper.

## References

1. Carrara E, Pfeffer I, Zusman O, Leibovici L, Paul M. Determinants of inappropriate empirical antibiotic treatment: systematic review and meta-analysis. Int J Antimicrob Agents. 2018 Apr;51(4):548–53. DOI: https://doi.org/10.1016/j.ijantimicag.2017.12.013

2. Chatterjee A, Modarai M, Naylor NR, Boyd SE, Atun R, Barlow J, et al. Quantifying drivers of antibiotic resistance in humans: a systematic review. Lancet Infect Dis. 2018 Dec;18(12):e368.#x2013;78. DOI: https://doi.org/10.1016/S1473-3099(18)30296-2

3. Holmes AH, Moore LSP, Sundsfjord A, Steinbakk M, Regmi S, Karkey A, et al. Understanding the mechanisms and drivers of antimicrobial resistance. The Lancet. 2016 Jan;387(10014):176–87. DOI: https://doi.org/10.1016/S0140-6736(15)00473-0

4. Bakhit M, Hoffmann T, Scott AM, Beller E, Rathbone J, Del Mar C. Resistance decay in individuals after antibiotic exposure in primary care: a systematic review and meta-analysis. BMC Med. 2018 Dec;16(1):126. DOI: https://doi.org/10.1186/s12916-018-1109-4

5. Bassetti M, Righi E, Vena A, Graziano E, Russo A, Peghin M. Risk stratification and treatment of ICU-acquired pneumonia caused by multidrug-resistant/extensively drug-resistant/pandrug-resistant bacteria: Curr Opin Crit Care. 2018 Oct;24(5):385–93. DOI: 10.1097/MCC.0000000000000534

6. Raman G, Avendano EE, Chan J, Merchant S, Puzniak L. Risk factors for hospitalized patients with resistant or multidrug-resistant Pseudomonas aeruginosa infections: a systematic review and meta-analysis. Antimicrob Resist Infect Control. 2018 Dec;7(1):79. DOI: https://doi.org/10.1186/s13756-018-0370-9

7. Prina E, Ranzani OT, Polverino E, Cillóniz C, Ferrer M, Fernandez L, et al. Risk Factors Associated with Potentially Antibiotic-Resistant Pathogens in Community-Acquired Pneumonia. Ann Am Thorac Soc. 2015 Feb;12(2):153–60. DOI: https://doi.org/10.1513/AnnalsATS.201407-305OC

8. Sulis G, Sayood S, Katukoori S, Bollam N, George I, Yaeger LH, et al. Exposure to World Health Organization’s AWaRe antibiotics and isolation of multidrug resistant bacteria: a systematic review and meta-analysis. Clin Microbiol Infect. 2022 Sep;28(9):1193–202. DOI: https://doi.org/10.1016/j.cmi.2022.03.014

9. Minh NNQ, Toi PV, Qui LM, Tinh LBB, Ngoc NT, Kim LTN, et al. Antibiotic use and prescription and its effects on Enterobacteriaceae in the gut in children with mild respiratory infections in Ho Chi Minh City, Vietnam. A prospective observational outpatient study. Foster J, editor. PLOS ONE. 2020 Nov 4;15(11):e0241760. DOI: https://doi.org/10.1371/journal.pone.0241760

10. Dan S, Shah A, Justo JA, Bookstaver PB, Kohn J, Albrecht H, et al. Prediction of Fluoroquinolone Resistance in Gram-Negative Bacteria Causing Bloodstream Infections. Antimicrob Agents Chemother. 2016 Apr;60(4):2265–72. DOI: https://doi.org/10.1128/AAC.02728-15

11. Shah A, Justo JA, Bookstaver PB, Kohn J, Albrecht H, Al-Hasan MN. Application of Fluoroquinolone Resistance Score in Management of Complicated Urinary Tract Infections. Antimicrob Agents Chemother. 2017 May;61(5):e02313-16. DOI: https://doi.org/10.1128/AAC.02313-16

12. Hastie T, Tibshirani R. Varying-Coefficient Models. J R Stat Soc Ser B Methodol. 1993 Sep;55(4):757–79. DOI: https://doi.org/10.1111/j.2517-6161.1993.tb01939.x

13. Cherny SS, Nevo D, Baraz A, Baruch S, Lewin-Epstein O, Stein GY, et al. Revealing antibiotic cross-resistance patterns in hospitalized patients through Bayesian network modelling. :10. DOI: https://doi.org/10.1093/jac/dkaa408

14. Chowers M, Zehavi T, Gottesman BS, Baraz A, Nevo D, Obolski U. Estimating the impact of cefuroxime versus cefazolin and amoxicillin/clavulanate use on future collateral resistance: a retrospective comparison. J Antimicrob Chemother. 2022 Apr 15;dkac130. DOI: https://doi.org/10.1093/jac/dkac130

