## Supplementary Information for "The time-varying association between previous antibiotic use and antibiotic resistance"

**Contents**

1. Additional information:
   a. Further details regarding the software and statistical methods employed.
   b. The antimicrobial susceptibility testing procedure.
   c. Key covariates distributions stratified by antibiotic resistance (Tables S1-S3).
   d. Descriptive statistics about antibiotic susceptibility tests stratified by bacterial species and sample sources (Tables S4-S6).
   e. Multivariable regression coefficients (Tables S7-S9).
2. Sensitivity analyses:
   a. Additional covariate adjustment for previous antibiotic use (Figures S1-S3).
   b. Additional adjustment for prior hospitalization duration (Figure S4).
   c. Subgroup analyses by bacterial species and sample sources (Figures S5-S7).
3. Exponential decay estimates: Approximations to the time-varying estimates by exponential rate of decay (Figure S8).

### Additional Information

### Software and specifications

All analyses were performed using the R v3.6.1, with the packages *mgcv* v1.8.28 [1] and *tableone* v0.10.0. The time-varying coefficients were modeled as cubic splines with 10 equi-spaced knots, and the model was estimated with Restricted Maximum Likelihood (REML). The null hypothesis that time-varying coefficients were equal to 0 was tested using a Wald-type test [1,2].

### Antibiotic susceptibility

Susceptibility test results were obtained as follows: for gram-negative bacteria in urine or wound cultures, the VITEK 2 system (bioMerieux, Durham, NC) was used. For gram-positive bacteria in urine and wounds, or any bacteria from blood cultures, disk diffusion tests with CLSI breakpoints were used. Nonsusceptible test outcomes (both intermediate and resistant) were aggregated into resistant results.

### Covariates distribution

|  |  | Susceptible | Resistant |
| --- | --- | --- | --- |
| *N* | (%) | 11054 (84) | 2040 (16) |
| *Age* | Mean (SD) | 69.43 (20.37) | 70.26 (18.19) |
| *Male* | N (%) | 5183 (46.9) | 1097 (53.8) |
| *Used aminoglycosides past year* | N (%) | 587 (5.3) | 281 (13.8) |
| *Had catheter* | N (%) | 2702 (24.4) | 789 (38.7) |
| *Nosocomial* | N (%) | 4306 (39) | 915 (44.9) |
| *Immunosuppression* | N (%) | 452 (4.1) | 95 (4.7) |
| *Diabetes* | N (%) | 1847 (16.7) | 407 (20.0) |
| *Arrived from home* | N (%) | 6856 (62.0) | 1057 (51.8) |
| *Arrived from an institution* | N (%) | 1391 (12.6) | 482 (23.6) |
| *Arrived from another hospital* | N (%) | 278 (2.5) | 82 (4.0) |
| *Arrived from: other* | N (%) | 898 (8.1) | 169 (8.3) |
| *Arrived from: unknown* | N (%) | 1631 (14.8) | 250 (12.3) |
| *# Hospitalizations past year* | Mean (SD) | 2.06 (1.52) | 2.60 (1.96) |
| *Had MRSA* | N (%) | 591 (5.3) | 255 (12.5) |
| *Had ESBL* | N (%) | 1678 (15.2) | 757 (37.1) |
| *Had other resistant infection* | N (%) | 410 (3.7) | 193 (9.5) |
| *Culture drawn at the internal unit* | N (%) | 5224 (47.3) | 1073 (52.6) |
| *Culture drawn at ER* | N (%) | 1099 (9.9) | 179 (8.8) |
| *Culture drawn at the orthopedic unit* | N (%) | 1178 (10.7) | 257 (12.6) |
| *Culture drawn at other unit* | N (%) | 812 (7.3) | 100 (4.9) |
| *Sample location: urine* | N (%) | 4688 (42.4) | 950 (46.6) |
| *Sample location: blood* | N (%) | 1329 (12.0) | 220 (10.8) |
| *Sample location: wound* | N (%) | 2431 (22.0) | 501 (24.6) |
| *Sample location: sputum* | N (%) | 833 (7.5) | 117 (5.7) |
| *Sample location: unknown* | N (%) | 1635 (14.8) | 222 (10.9) |
| *Sample location: other* | N (%) | 138 (1.2) | 30 (1.5) |
| *Genus: Escherichia* | N (%) | 4437 (40.1) | 698 (34.2) |
| *Genus: Enterobacter* | N (%) | 460 (4.2) | 32 (1.6) |
| *Genus: Citrobacter* | N (%) | 494 (4.5) | 15 (0.7) |
| *Genus: Pseudomonas* | N (%) | 2095 (19.0) | 307 (15.0) |
| *Genus: Proteus* | N (%) | 937 (8.5) | 273 (13.4) |
| *Genus: Klebsiella* | N (%) | 1605 (14.5) | 421 (20.6) |
| *Genus: Morganella* | N (%) | 312 ( 2.8) | 51 ( 2.5) |
| *Genus: other* | N (%) | 714 ( 6.5) | 243 (11.9) |

**Table S1.** **Summary of key covariates used in this study, stratified by gentamicin resistance result.** #: number of. MRSA: Methicillin-resistant *Staphylococcus aureus*. ESBL: extended spectrum beta-lactamase.

|  |  | Susceptible | Resistant |
| --- | --- | --- | --- |
| *N* | (%) | 9883 (76) | 3167 (24) |
| *Age* | Mean (SD) | 68.86 (20.41) | 72.68 (17.22) |
| *Male* | N (%) | 4720 (47.8) | 1527 (48.2) |
| *Used cephalosporins past year* | N (%) | 3829 (38.7) | 1666 (52.6) |
| *Used non-cephalosporin beta-lactams past year* | N (%) | 1717 (17.4) | 756 (23.9) |
| *Had catheter* | N (%) | 2362 (23.9) | 1154 (36.4) |
| *Nosocomial* | N (%) | 3885 (39.3) | 1343 (42.4) |
| *Immunosuppression* | N (%) | 411 (4.2) | 139 (4.4) |
| *Diabetes* | N (%) | 1628 (16.5) | 643 (20.3) |
| *Arrived from home* | N (%) | 6269 (63.4) | 1607 (50.7) |
| *Arrived from an institution* | N (%) | 1122 (11.4) | 764 (24.1) |
| *Arrived from another hospital* | N (%) | 234 (2.4) | 122 (3.9) |
| *Arrived from: other* | N (%) | 799 (8.1) | 269 (8.5) |
| *Arrived from: unknown* | N (%) | 1459 (14.8) | 405 (12.8) |
| *# Hospitalizations past year* | Mean (SD) | 2.03 (1.51) | 2.52 (1.83) |
| *Had MRSA* | N (%) | 544 (5.5) | 310 (9.8) |
| *Had ESBL* | N (%) | 1020 (10.3) | 1427 (45.1) |
| *Had other resistant infection* | N (%) | 381 (3.9) | 231 (7.3) |
| *Culture drawn at the internal unit* | N (%) | 4508 (45.6) | 1761 (55.6) |
| *Culture drawn at ER* | N (%) | 971 (9.8) | 309 (9.8) |
| *Culture drawn at the orthopedic unit* | N (%) | 1195 (12.1) | 264 (8.3) |
| *Culture drawn at other unit* | N (%) | 718 (7.3) | 157 (5.0) |
| *Sample location: urine* | N (%) | 4024 (40.7) | 1668 (52.7) |
| *Sample location: blood* | N (%) | 1149 (11.6) | 336 (10.6) |
| *Sample location: wound* | N (%) | 2371 (24.0) | 590 (18.6) |
| *Sample location: sputum* | N (%) | 773 (7.8) | 204 (6.4) |
| *Sample location: unknown* | N (%) | 1486 (15.0) | 366 (11.6) |
| *Sample location: other* | N (%) | 80 (0.8) | 3 (0.1) |
| *Genus: Escherichia* | N (%) | 3637 (36.8) | 1528 (48.2) |
| *Genus: Enterobacter* | N (%) | 381 (3.9) | 109 (3.4) |
| *Genus: Citrobacter* | N (%) | 460 (4.7) | 50 (1.6) |
| *Genus: Pseudomonas* | N (%) | 2229 (22.6) | 188 (5.9) |
| *Genus: Proteus* | N (%) | 943 (9.5) | 313 (9.9) |
| *Genus: Klebsiella* | N (%) | 1251 (12.7) | 779 (24.6) |
| *Genus: Morganella* | N (%) | 337 (3.4) | 34 (1.1) |
| *Genus: other* | N (%) | 645 (6.5) | 166 (5.2) |

**Table S2.** **Summary of key covariates used in this study, stratified by ceftazidime resistance result.** #: number of. MRSA: Methicillin-resistant *Staphylococcus aureus*. ESBL: extended spectrum beta-lactamase.

|  |  | Susceptible | Resistant |
| --- | --- | --- | --- |
| *N* | (%) | 11267 (73) | 4095 (27) |
| *Age* | Mean (SD) | 67.55 (21.09) | 72.67 (16.97) |
| *Male* | N (%) | 5511 (48.9) | 2223 (54.3) |
| *Used fluoroquinolones past year* | N (%) | 407 (3.6) | 435 (10.6) |
| *Had catheter* | N (%) | 2472 (21.9) | 1487 (36.3) |
| *Nosocomial* | N (%) | 4446 (39.5) | 1518 (37.1) |
| *Immunosuppression* | N (%) | 466 (4.1) | 192 (4.7) |
| *Diabetes* | N (%) | 1772 (15.7) | 904 (22.1) |
| *Arrived from home* | N (%) | 7237 (64.2) | 1982 (48.4) |
| *Arrived from an institution* | N (%) | 1226 (10.9) | 1133 (27.7) |
| *Arrived from another hospital* | N (%) | 256 (2.3) | 168 (4.1) |
| *Arrived from: other* | N (%) | 939 (8.3) | 362 (8.8) |
| *Arrived from: unknown* | N (%) | 1609 (14.3) | 450 (11.0) |
| *# Hospitalizations past year* | Mean (SD) | 2.01 (1.47) | 2.53 (1.87) |
| *Had MRSA* | N (%) | 556 (4.9) | 882 (21.5) |
| *Had ESBL* | N (%) | 1418 (12.6) | 1189 (29.0) |
| *Had other resistant infection* | N (%) | 385 (3.4) | 336 (8.2) |
| *Culture drawn at the internal unit* | N (%) | 5126 (45.5) | 2329 (56.9) |
| *Culture drawn at ER* | N (%) | 1034 (9.2) | 340 (8.3) |
| *Culture drawn at the orthopedic unit* | N (%) | 1501 (13.3) | 537 (13.1) |
| *Culture drawn at other unit* | N (%) | 856 (7.6) | 173 (4.2) |
| *Sample location: urine* | N (%) | 3950 (35.1) | 1607 (39.2) |
| *Sample location: blood* | N (%) | 1446 (12.8) | 374 (9.1) |
| *Sample location: wound* | N (%) | 2873 (25.5) | 877 (21.4) |
| *Sample location: sputum* | N (%) | 926 (8.2) | 253 (6.2) |
| *Sample location: unknown* | N (%) | 1872 (16.6) | 944 (23.1) |
| *Sample location: other* | N (%) | 200 (1.8) | 40 (1.0) |
| *Genus: Escherichia* | N (%) | 3533 (31.4) | 1545 (37.7) |
| *Genus: Enterobacter* | N (%) | 450 (4.0) | 35 (0.9) |
| *Genus: Citrobacter* | N (%) | 494 (4.4) | 16 (0.4) |
| *Genus: Pseudomonas* | N (%) | 2074 (18.4) | 347 (8.5) |
| *Genus: Proteus* | N (%) | 696 (6.2) | 456 (11.1) |
| *Genus: Klebsiella* | N (%) | 1557 (13.8) | 327 (8.0) |
| *Genus: Morganella* | N (%) | 219 (1.9) | 115 (2.8) |
| *Genus: other* | N (%) | 826 (7.3) | 226 (5.5) |

**Table S3. Summary of key covariates used in this study, stratified by ciprofloxacin resistance result.** #: number of. MRSA: Methicillin-resistant *Staphylococcus aureus*. ESBL: extended spectrum beta-lactamase.

| **Bacterial species** | **Urine** | **Blood** | **Wound** | **Sputum** | **Other** | **Unknown** | **Total** |
| --- | --- | --- | --- | --- | --- | --- | --- |
| *Escherichia coli* | 2828 | 782 | 649 | 60 | 23 | 786 | 5128 |
| *Pseudomonas aeruginosa* | 864 | 117 | 625 | 490 | 35 | 223 | 2354 |
| *Klebsiella pneumoniae* | 966 | 203 | 260 | 128 | 4 | 260 | 1821 |
| *Proteus mirabilis* | 436 | 127 | 393 | 40 | 1 | 124 | 1121 |
| *Enterobacter cloacae* | 93 | 32 | 207 | 43 | 2 | 97 | 474 |
| *Morganella morganii* | 50 | 25 | 227 | 7 | 1 | 53 | 363 |
| *Citrobacter koseri* | 117 | 22 | 127 | 23 | 2 | 40 | 331 |
| *Klebsiella oxytoca* | 47 | 26 | 60 | 13 | 0 | 50 | 196 |
| *Serratia marcescens* | 33 | 22 | 56 | 53 | 5 | 21 | 190 |
| *Enterobacter aerogenes* | 56 | 10 | 25 | 28 | 3 | 37 | 159 |
| *Citrobacter freundii* | 31 | 7 | 65 | 6 | 2 | 32 | 143 |
| *Providencia stuartii* | 43 | 9 | 40 | 6 | 0 | 16 | 114 |
| *Acinetobacter baumannii* | 18 | 9 | 27 | 35 | 0 | 19 | 108 |
| Other | 56 | 158 | 171 | 19 | 90 | 99 | 593 |
|  | 5638 | 1549 | 2932 | 951 | 168 | 1857 | 13095 |

**Table S4**. **Gentamicin susceptibility tests stratified by bacterial species and sample sources.** Bacterial species with $n<100$ samples were aggregated into "other", as were rare sample sources (including ear, eye, stool, rectal, sinus, vaginal, cervix, throat and csf).

| **Bacterial species** | **Urine** | **Blood** | **Wound** | **Sputum** | **Other** | **Unknown** | **Total** |
| --- | --- | --- | --- | --- | --- | --- | --- |
| *Escherichia coli* | 2855 | 779 | 659 | 61 | 23 | 781 | 5158 |
| *Pseudomonas aeruginosa* | 873 | 115 | 616 | 493 | 36 | 236 | 2369 |
| *Klebsiella pneumoniae* | 960 | 205 | 256 | 125 | 4 | 275 | 1825 |
| *Proteus mirabilis* | 456 | 130 | 409 | 42 | 1 | 127 | 1165 |
| *Enterobacter cloacae* | 91 | 32 | 205 | 41 | 2 | 103 | 474 |
| *Morganella morganii* | 49 | 30 | 239 | 7 | 1 | 45 | 371 |
| *Citrobacter koseri* | 121 | 25 | 129 | 23 | 3 | 31 | 332 |
| *Klebsiella oxytoca* | 46 | 25 | 63 | 13 | 0 | 49 | 196 |
| *Serratia marcescens* | 32 | 22 | 55 | 55 | 5 | 21 | 190 |
| *Enterobacter aerogenes* | 53 | 7 | 25 | 28 | 2 | 37 | 152 |
| *Citrobacter freundii* | 32 | 7 | 62 | 6 | 1 | 35 | 143 |
| *Acinetobacter baumannii* | 17 | 10 | 31 | 47 | 0 | 19 | 124 |
| *Providencia stuartii* | 45 | 9 | 39 | 6 | 1 | 14 | 114 |
| Other | 62 | 89 | 173 | 31 | 4 | 79 | 438 |
|  | 5692 | 1485 | 2961 | 978 | 83 | 1852 | 13051 |

**Table S5. Ceftazidime susceptibility tests stratified by bacterial species and sample sources.** Bacterial species with $n<100$ samples were aggregated into "other", as were rare sample sources (including ear, eye, stool, rectal, sinus, vaginal, cervix, throat and csf).

| **Bacterial species** | **Urine** | **Blood** | **Wound** | **Sputum** | **Other** | **Unknown** | **Total** |
| --- | --- | --- | --- | --- | --- | --- | --- |
| *Escherichia coli* | 2794 | 783 | 639 | 59 | 23 | 773 | 5071 |
| *Pseudomonas aeruginosa* | 882 | 116 | 612 | 495 | 37 | 231 | 2373 |
| *Staphylococcus aureus* | 34 | 223 | 792 | 148 | 30 | 908 | 2139 |
| *Klebsiella pneumoniae* | 891 | 197 | 234 | 118 | 4 | 237 | 1681 |
| *Proteus mirabilis* | 412 | 119 | 383 | 38 | 1 | 109 | 1062 |
| *Enterobacter cloacae* | 88 | 32 | 205 | 42 | 2 | 100 | 469 |
| *Morganella morganii* | 45 | 25 | 207 | 7 | 2 | 48 | 334 |
| *Citrobacter koseri* | 120 | 25 | 126 | 22 | 3 | 34 | 330 |
| *Klebsiella oxytoca* | 47 | 27 | 59 | 13 | 0 | 48 | 194 |
| *Serratia marcescens* | 30 | 23 | 55 | 55 | 5 | 20 | 188 |
| *Enterobacter aerogenes* | 57 | 12 | 25 | 27 | 3 | 32 | 156 |
| *Citrobacter freundii* | 32 | 7 | 63 | 6 | 1 | 36 | 145 |
| *Acinetobacter baumannii* | 17 | 10 | 32 | 47 | 0 | 19 | 125 |
| *Staphylococcus coagulase negative* | 0 | 63 | 28 | 0 | 1 | 31 | 123 |
| *Haemophilus influenzae* | 0 | 13 | 6 | 62 | 18 | 23 | 123 |
| *Providencia stuartii* | 41 | 7 | 35 | 6 | 0 | 15 | 104 |
| Other | 67 | 138 | 249 | 36 | 105 | 152 | 747 |
|  | 5557 | 1820 | 3750 | 1181 | 235 | 2816 | 15364 |

**Table S6. Ciprofloxacin susceptibility tests stratified by bacterial species and sample sources.** Bacterial species with $n<100$ samples were aggregated into "other", as were rare sample sources (including ear, eye, stool, rectal, sinus, vaginal, cervix, throat and csf).

### Regression coefficients

| **Resistance to gentamicin** | | | |
| --- | --- | --- | --- |
| *Variable* | *Odds Ratio* | *95% CI* | *P-value* |
| Age | 1.00 | 0.99 – 1.00 | 0.538 |
| Sex (male) | 1.31 | 1.12 – 1.52 | <0.001 |
| **#** Times hospitalized past year | 1.05 | 1.00 – 1.10 | 0.033 |
| Had catheter | 1.29 | 1.08 – 1.54 | 0.004 |
| Nosocomial | 1.23 | 1.06 – 1.42 | 0.005 |
| Diabetes | 1.00 | 0.82 – 1.21 | 0.976 |
| Immunosuppression | 1.03 | 0.72 – 1.48 | 0.859 |
| Culture drawn at the surgical unit | 0.96 | 0.78 – 1.17 | 0.660 |
| Culture drawn at ER | 0.97 | 0.60 – 1.55 | 0.889 |
| Culture drawn at the orthopedic unit | 1.18 | 0.89 – 1.56 | 0.239 |
| Culture drawn at other unit | 0.61 | 0.42 – 0.89 | 0.010 |
| Sample source: sputum | 0.43 | 0.32 – 0.57 | <0.001 |
| Sample source: blood | 0.75 | 0.61 – 0.92 | 0.005 |
| Sample source: wound | 0.88 | 0.72 – 1.09 | 0.238 |
| Sample source: unknown | 0.68 | 0.55 – 0.85 | 0.001 |
| Sample source: other | 1.13 | 0.65 – 1.98 | 0.668 |
| Genus: Enterobacter | 0.54 | 0.35 – 0.83 | 0.005 |
| Genus: Citrobacter | 0.20 | 0.11 – 0.35 | <0.001 |
| Genus: Pseudomonas | 0.81 | 0.67 – 0.99 | 0.037 |
| Genus: Proteus | 1.33 | 1.08 – 1.65 | 0.008 |
| Genus: Klebsiella | 1.61 | 1.35 – 1.92 | <0.001 |
| Genus: Morganella | 0.85 | 0.58 – 1.26 | 0.420 |
| Genus: other | 2.61 | 2.07 – 3.30 | <0.001 |
| Arrived from an institution | 1.84 | 1.51 – 2.24 | <0.001 |
| Arrived from another hospital | 1.76 | 1.16 – 2.69 | 0.008 |
| Arrived from: unknown | 1.62 | 1.05 – 2.49 | 0.029 |
| Arrived from: other | 1.02 | 0.78 – 1.34 | 0.895 |
| Had ESBL | 3.29 | 2.78 – 3.90 | <0.001 |
| Had MRSA | 1.44 | 1.09 – 1.89 | 0.009 |
| Had other resistant infection | 1.58 | 1.14 – 2.20 | 0.006 |
| S(Days since aminoglycosides use) |  |  | <0.001 |
| Observations | 13094 | | |

**Table S7**. **Estimates of the mixed-effects time-varying logistic models fitted to resistance to gentamicin as outcome**. #: number of. MRSA: Methicillin-resistant *Staphylococcus aureus*. ESBL: extended spectrum beta-lactamase. S(Days since aminoglycosides use): The time-varying coefficient of *Used aminoglycosides,* testing if this coefficient equals zero for all *t*.

| **Resistance to ceftazidime** | | | |
| --- | --- | --- | --- |
| *Variable* | *Odds Ratio* | *95% CI* | *P-value* |
| Age | 1.01 | 1.00 – 1.01 | 0.005 |
| Sex (male) | 1.16 | 1.01 – 1.33 | 0.031 |
| Used non-caphalosporin beta-lactams | 1.24 | 1.01 – 1.50 | 0.036 |
| Used non-caphalosporin beta-lactams*  Days since cephalosporins use | 1 | 1.00 – 1.00 | 0.468 |
| **#** Times hospitalized past year | 1.04 | 0.99 – 1.09 | 0.163 |
| Had catheter | 1.11 | 0.94 – 1.30 | 0.225 |
| Nosocomial | 1.33 | 1.15 – 1.53 | <0.001 |
| Diabetes | 1.19 | 1.00 – 1.41 | 0.053 |
| Immunosuppression | 1.05 | 0.76 – 1.44 | 0.768 |
| Culture drawn at the surgical unit | 0.82 | 0.69 – 0.98 | 0.031 |
| Culture drawn at ER | 1.34 | 0.87 – 2.06 | 0.183 |
| Culture drawn at the orthopedic unit | 0.91 | 0.69 – 1.19 | 0.471 |
| Culture drawn at other unit | 0.64 | 0.46 – 0.89 | 0.009 |
| Sample source: sputum | 0.72 | 0.56 – 0.94 | 0.014 |
| Sample source: blood | 0.59 | 0.49 – 0.71 | <0.001 |
| Sample source: wound | 0.7 | 0.57 – 0.85 | <0.001 |
| Sample source: unknown | 0.66 | 0.54 – 0.80 | <0.001 |
| Sample source: other | 0.36 | 0.10 – 1.33 | 0.124 |
| Genus: Enterobacter | 0.79 | 0.59 – 1.06 | 0.12 |
| Genus: Citrobacter | 0.28 | 0.20 – 0.41 | <0.001 |
| Genus: Pseudomonas | 0.1 | 0.08 – 0.13 | <0.001 |
| Genus: Proteus | 0.45 | 0.37 – 0.55 | <0.001 |
| Genus: Klebsiella | 1.37 | 1.17 – 1.59 | <0.001 |
| Genus: Morganella | 0.16 | 0.10 – 0.25 | <0.001 |
| Genus: other | 0.5 | 0.39 – 0.64 | <0.001 |
| Arrived from an institution | 2.23 | 1.86 – 2.67 | <0.001 |
| Arrived from another hospital | 2.07 | 1.40 – 3.06 | <0.001 |
| Arrived from: unknown | 1.28 | 0.86 – 1.91 | 0.221 |
| Arrived from: other | 1.38 | 1.08 – 1.76 | 0.01 |
| Had ESBL | 9.21 | 7.87 – 10.78 | <0.001 |
| Had MRSA | 1.09 | 0.83 – 1.42 | 0.554 |
| Had other resistant infection | 1.64 | 1.20 – 2.25 | 0.002 |
| S(Days since cephalosporins use) |  |  | <0.001 |
| Observations | 13050 | | |

**Table S8**. **Estimates of the mixed-effects time-varying logistic models fitted to resistance to ceftazidime as outcome**. *: Interaction term between *Used non-cephalosporin beta-lactams* and *Days since cephalosporins use*. #: number of. MRSA: Methicillin-resistant Staphylococcus aureus. ESBL: extended spectrum beta-lactamase. S(Days since cephalosporins use): The time-varying coefficient of *Used cephalosporins*, testing if this coefficient equals zero for all *t*.

| **Resistance to ciprofloxacin** | | | |
| --- | --- | --- | --- |
| *Variable* | *Odds Ratio* | *95% CI* | *P-value* |
| Age | 1.01 | 1.01 – 1.02 | <0.001 |
| Sex (male) | 1.33 | 1.18 – 1.50 | <0.001 |
| **#** Times hospitalized past year | 1.05 | 1.02 – 1.10 | 0.006 |
| Had catheter | 1.41 | 1.23 – 1.62 | <0.001 |
| Nosocomial | 1.03 | 0.92 – 1.16 | 0.619 |
| Diabetes | 1.29 | 1.10 – 1.50 | 0.001 |
| Immunosuppression | 1.06 | 0.80 – 1.39 | 0.704 |
| Culture drawn at the surgical unit | 0.91 | 0.78 – 1.07 | 0.268 |
| Culture drawn at the orthopedic unit | 0.84 | 0.68 – 1.04 | 0.112 |
| Culture drawn at ER | 1.02 | 0.70 – 1.48 | 0.911 |
| Culture drawn at other unit | 0.59 | 0.44 – 0.79 | <0.001 |
| Sample source: blood | 0.47 | 0.40 – 0.56 | <0.001 |
| Sample source: wound | 0.64 | 0.53 – 0.76 | <0.001 |
| Sample source: sputum | 0.53 | 0.42 – 0.67 | <0.001 |
| Sample source: unknown | 0.94 | 0.80 – 1.10 | 0.424 |
| Sample source: other | 1.33 | 0.84 – 2.09 | 0.226 |
| Genus: Enterobacter | 0.16 | 0.11 – 0.24 | <0.001 |
| Genus: Citrobacter | 0.07 | 0.04 – 0.11 | <0.001 |
| Genus: Staphylococcus | 1.57 | 1.33 – 1.85 | <0.001 |
| Genus: Pseudomonas | 0.21 | 0.18 – 0.26 | <0.001 |
| Genus: Proteus | 1.14 | 0.95 – 1.37 | 0.159 |
| Genus: Klebsiella | 0.34 | 0.28 – 0.40 | <0.001 |
| Genus: Morganella | 0.94 | 0.69 – 1.29 | 0.712 |
| Genus: other | 0.55 | 0.44 – 0.68 | <0.001 |
| Arrived from an institution | 2.16 | 1.85 – 2.53 | <0.001 |
| Arrived from another hospital | 2.42 | 1.71 – 3.41 | <0.001 |
| Arrived from: unknown | 1.41 | 1.00 – 1.98 | 0.05 |
| Arrived from: other | 1.2 | 0.97 – 1.48 | 0.089 |
| Had ESBL | 2.54 | 2.19 – 2.95 | <0.001 |
| Had MRSA | 3.55 | 2.91 – 4.33 | <0.001 |
| Had other resistant infection | 2.1 | 1.58 – 2.79 | <0.001 |
| S(Days since fluoroquinolones use) |  |  | <0.001 |
| Observations | 15362 | | |

**Table S9**. **Estimates of the mixed-effects time-varying logistic models fitted to resistance to ciprofloxacin as outcome.** #: number of. MRSA: Methicillin-resistant *Staphylococcus aureus*. ESBL: extended spectrum beta-lactamase. S(Days since fluoroquinolones use): The time-varying coefficient of *Used fluoroquinolones,* testing if this coefficient equals zero for all *t*.

### Sensitivity analyses

#### Adjusting for additional antibiotic use

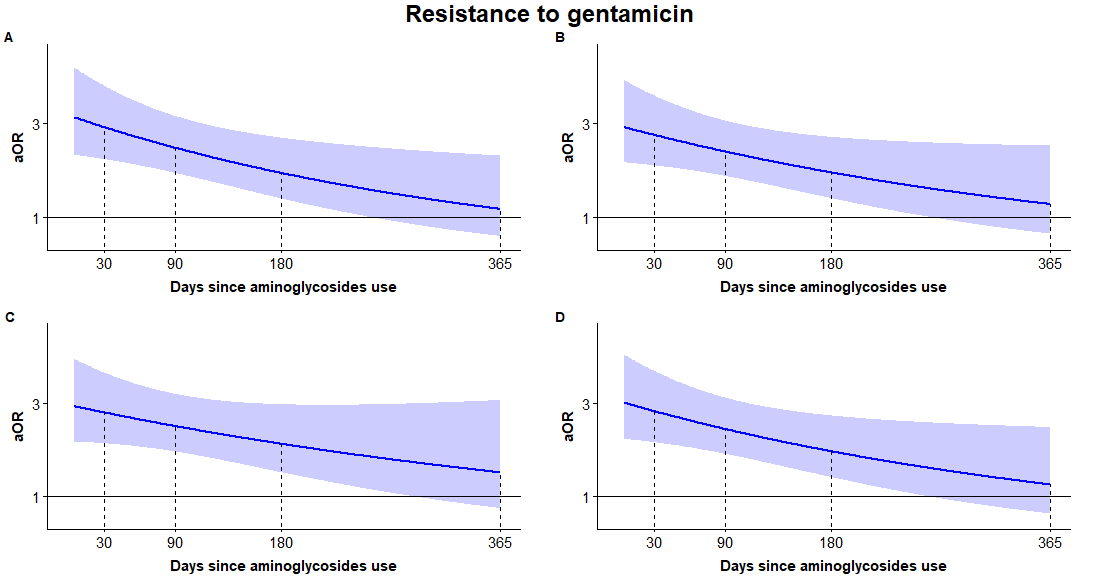

**Fig. S1. Estimates of risk for antibiotic resistance from mixed-effects time-varying logistic models:** Spline-based estimates (curves) and their 95% CI (shaded regions) of the aORs for gentamicin resistance, against days elapsed since last use of aminoglycosides. (a) no additional antibiotic use was controlled; (b) prior use of non-cephalosporin beta-lactams additionally controlled for; (c) prior use of cephalosporin additionally controlled for; and (d) prior use of fluoroquinolones additionally controlled for.

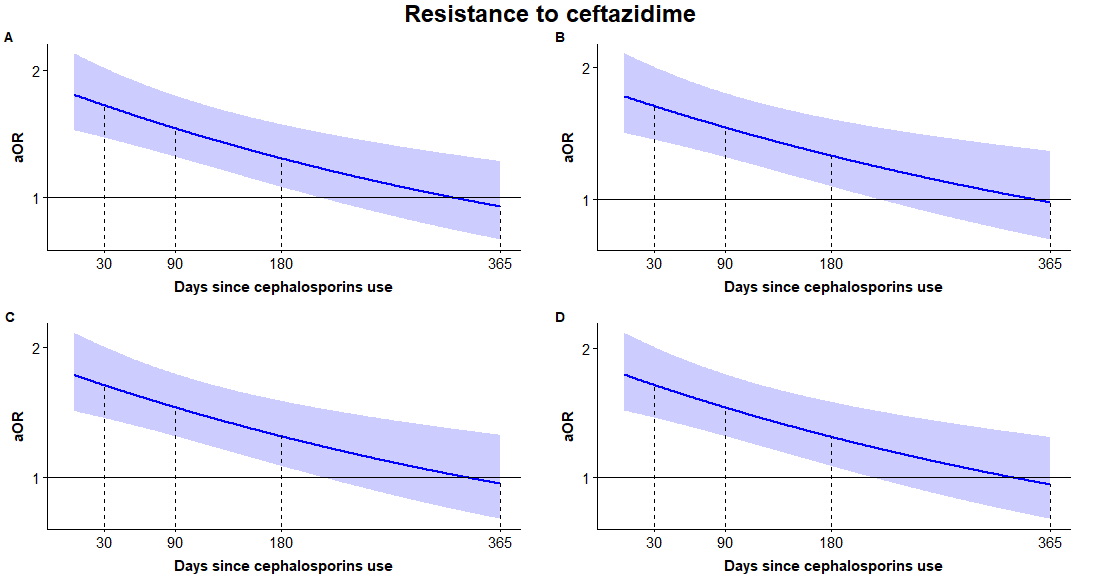

**Fig. S2. Estimates of risk for antibiotic resistance from mixed-effects time-varying logistic models:** Spline-based estimates (curves) and their 95% CI (shaded regions) of the aORs for ceftazidime resistance, against days elapsed since last use of cephalosporins. (a) no additional antibiotic use was controlled; (b) prior use of aminoglycosides additionally controlled for; (c) prior use of non-cephalosporin beta-lactams additionally controlled for; and (d) prior use of fluoroquinolones additionally controlled for.

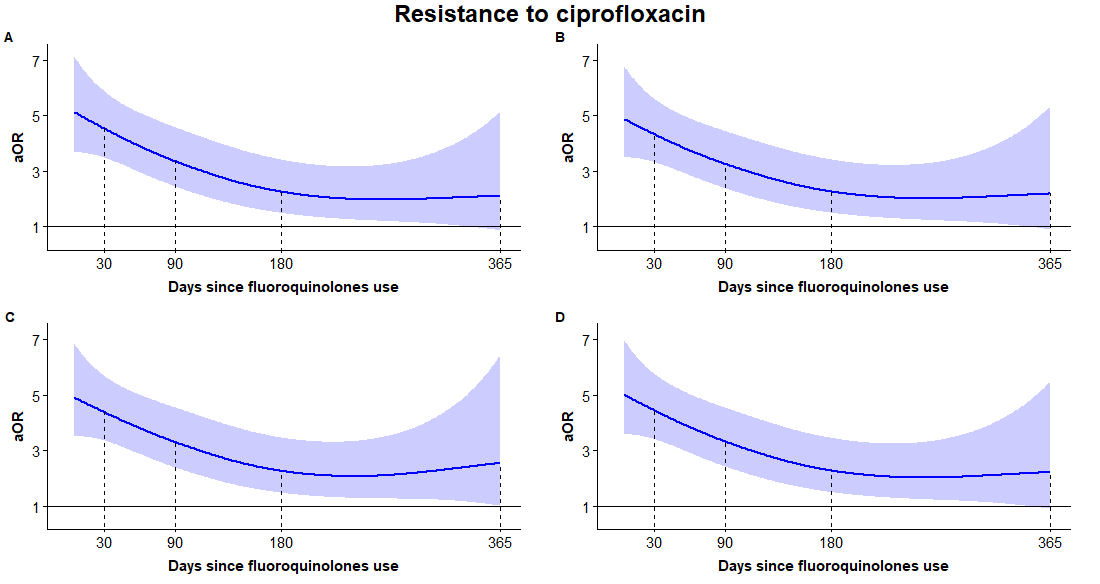

**Fig. S3. Estimates of risk for antibiotic resistance from mixed-effects time-varying logistic models:** Spline-based estimates (curves) and their 95% CI (shaded regions) of the aORs for ciprofloxacin resistance, against days elapsed since last use of fluoroquinolones. (a) no additional antibiotic use was controlled; (b) prior use of aminoglycosides additionally controlled for; (c) prior use of non-cephalosporin beta-lactams additionally controlled for; and (d) prior use of cephalosporins additionally controlled for.

#### Adjusting for prior hospitalization duration

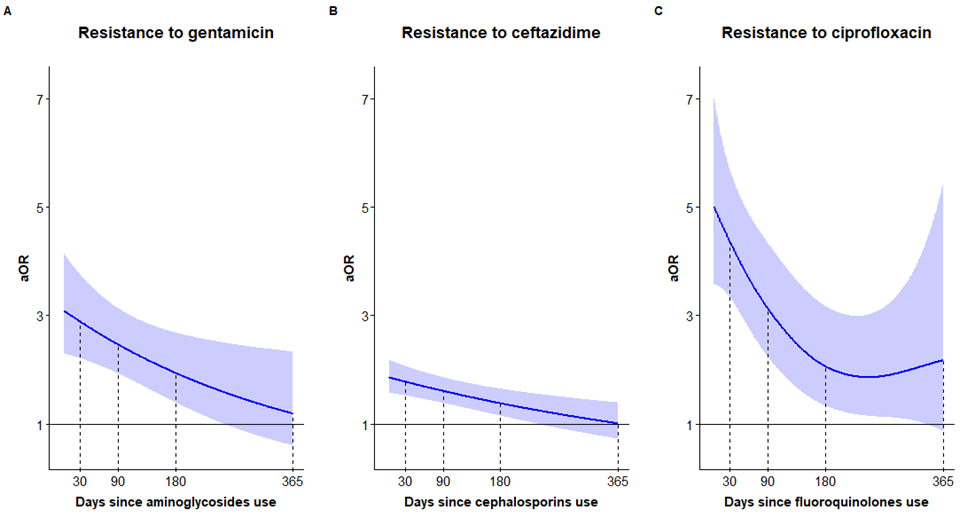

**Fig. S4. Sensitivity analysis of the estimates of risk for antibiotic resistance from mixed-effects time-varying logistic models, analogous to Figure 1 in the main text.** The multivariate model used here substitutes the number of times hospitalized in the prior year (Tables S7-S9) with the number of hospitalization days in the prior year. Results remained highly similar.

#### Subgroup analyses by common bacterial species and sample sources

For each antibiotic (gentamicin, ceftazidime and ciprofloxacin), subgroup analyses were performed by fitting the same model employed in the main text separately to common bacterial species (*Escherichia coli* and *Pseudomonas aeruginosa*) and common sample sources (urine and blood). Figures S5-S8 show the time-varying association between previous antibiotic use and resistance for these subgroups, for different antibiotics. The overall time-varying association patterns are retained. However, the subgroup analyses were by definition performed on a smaller sample size and hence the variance of the estimates is higher. Note that the models in Figure S5D and Figure S7B were fitted only up to 180 days due to data sparseness.

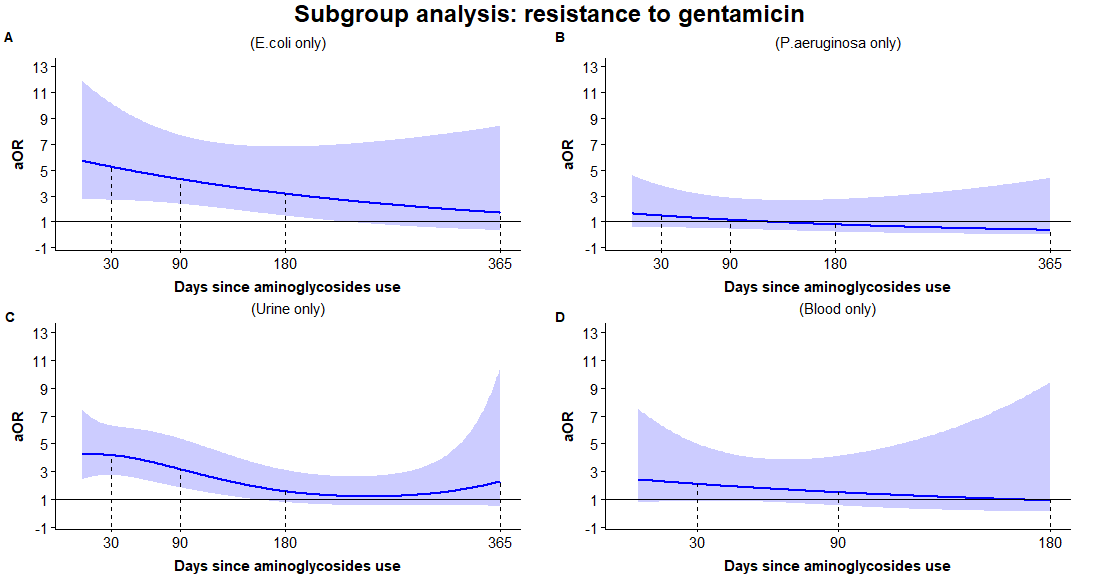

**Fig. S5. Subgroup analysis of gentamicin resistance by common bacterial species and sample locations:** Spline-based estimates (curves) and their 95% CI (shaded) of the aORs for resistance, against days elapsed since last use of aminoglycosides. (a) refers only to samples of *Escherichia coli* (n=5128); (b) refers only to samples of *Pseudomonas aeruginosa* (n=2354); (c) refers only to urine samples (n=5638); (d) refers only to blood samples (n=1537). Note that (d) refers to 180 days only, due to sparse data between day 180 and day 365 since last aminoglycoside use.

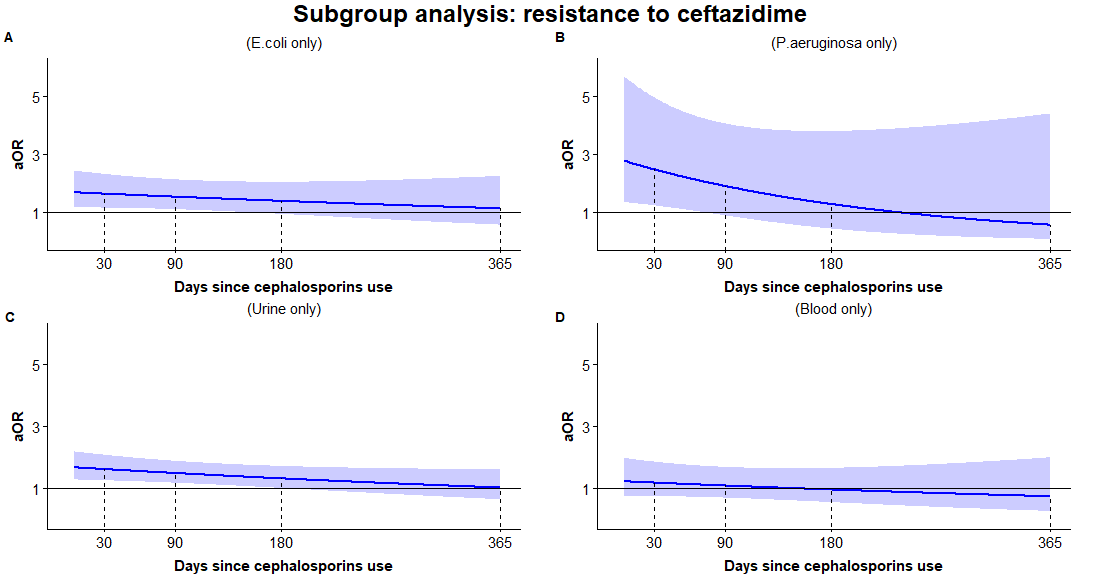

**Fig. S6. Subgroup analysis of ceftazidime resistance by common bacterial species and sample locations:** Spline-based estimates (curves) and their 95% CI (shaded) of the aORs for resistance, against days elapsed since last use of aminoglycosides. (a) refers only to samples of *Escherichia coli* (n=5158); (b) refers only to samples of *Pseudomonas aeruginosa* (n=2369); (c) refers only to urine samples (n=5962); (d) refers only to blood samples (n=1485)

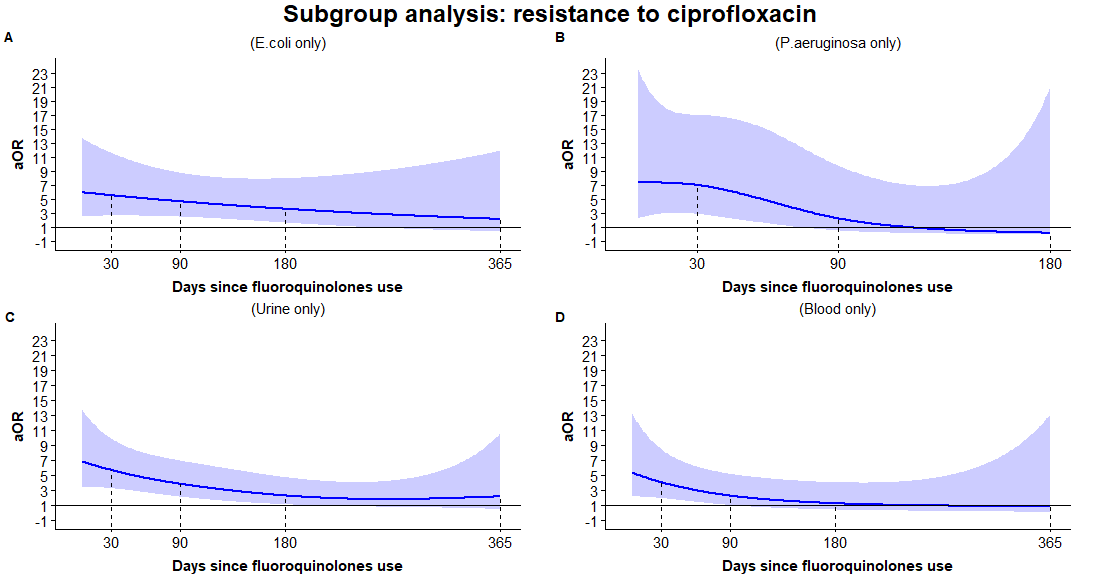

**Fig. S7. Subgroup analysis of gentamicin resistance by common bacterial species and sample locations:** Spline-based estimates (curves) and their 95% CI (shaded) of the aORs for resistance, against days elapsed since last use of aminoglycosides. (a) refers to data from *escherichia coli* only (n=5071); (b) refers to data from *pseudomonas aeruginosa* only (n=2335); (c) refers to data from urine cultures only (n=5557); (d) refers to data from blood cultures only (n=1820). Note that (b) refers to 180 days only, due to sparse data between day 180 and day 365 since last fluoroquinolone use.

### Exponential decay estimates

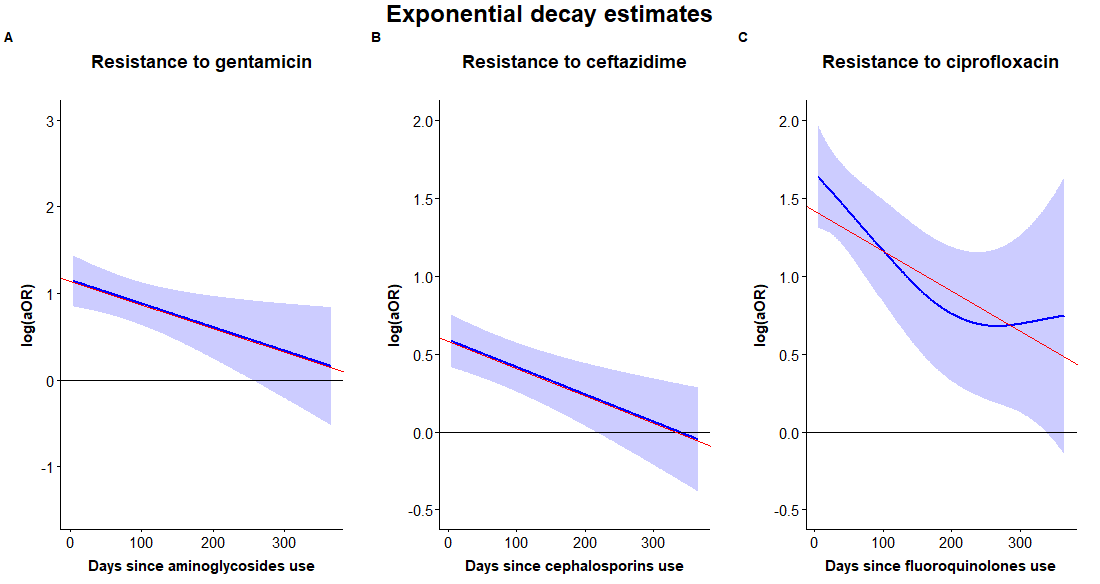

**Fig. S8. Estimating the exponential rate of decay: we use a linear regression (red) between the estimated log(aOR) and time to approximate the mixed-effects time-varying logistic models (blue).** (a) gentamicin resistance against last use of aminoglycosides was estimated as $log(aOR)=1.139528-0.002725t$; (b) ceftazidime resistance against last use of cephalosporins was estimated as $log(aOR)=0.575344-0.001655t$; and (c) ciprofloxacin resistance against last use of fluoroquinolones was estimated as $log(aOR)=1.1417894-0.002561t$, where $t$ is measured in days and transformed in the main text to months for convenience.

### References

1. Wood SN. Generalized additive models: an introduction with R. Second edition. Boca Raton: CRC Press/Taylor & Francis Group; 2017. 476 p. (Chapman & Hall/CRC texts in statistical science). DOI: <https://doi.org/10.1201/9781420010404>

2. Wood SN. On p-values for smooth components of an extended generalized additive model. Biometrika. 2013 Mar 1;100(1):221–8. DOI: <https://doi.org/10.1093/biomet/ass048>
